# Rare pathogenic variants in G-protein coupled receptor genes involved in gut-to-host communication are associated with cardiovascular disease risk

**DOI:** 10.1101/2024.10.01.24314734

**Authors:** Leticia Camargo Tavares, Rikeish R. Muralitharan, Matthew Snelson, Francine Z. Marques

**Author notes:** **Corresponding authors:** Dr Leticia Camargo Tavares, PhD, Postdoctoral Research Fellow, Hypertension Research Laboratory, School of Biological Sciences, Faculty of Science, Monash University, Melbourne, Australia.; Prof Francine Marques, Hypertension Research Laboratory, Victorian Heart Institute, Level 2, Victorian Heart Hospital, 631 Blackburn Road Clayton, VIC 3168 Monash University, Melbourne, Australia. **Data availability** This research has been conducted using the UK Biobank Resource under Application Number 86879. This work uses data provided by patients and collected by the NHS as part of their care and support. Full methods can be accessed at medRxiv: https://doi.org/10.1101/2024.10.01.24314734. **Author contributions** LCT led and FZM supported study design and conception; LCT led the formal data analysis and visualization; LCT led and RRM, MS, and FZM supported data interpretation; LCT led and RRM, MS, and FZM supported drafting the manuscript; all authors approved the final version of the manuscript.

## Abstract

**Background:** Gut microbial metabolites called short-chain fatty acids (SCFA) confer protective effects against cardiovascular disease and high blood pressure. Proposed mechanisms include anti-inflammatory signalling mediated by SCFA-sensing G-protein-coupled receptors (GPCR), particularly GPR41, GPR43, and GPR109a, as suggested by knockout mouse models. We aimed to determine if rare pathogenic variants (RPVs) affecting GPCR genes in humans increase the risk of hypertension (HTN) and major adverse cardiac events (MACEs), including acute coronary syndrome, heart failure, and ischemic stroke.

**Methods:** Using UK Biobank whole-exome sequencing data from 393,649 European participants, we identified rare (minor allele frequency <1%) pathogenic variants with predicted high-impact functional consequences in GPCR genes, based on Ensembl Variant Effect Predictor annotations. For missense variants, pathogenicity likelihood scores from AlphaMissense, Mendelian Clinically Applicable Pathogenicity, and Combined Annotation Dependent Depletion were assessed. Multivariable logistic regression models, adjusted for age, sex, BMI, genetic ancestry, and other potential confounders, were conducted to compare RPV prevalence between cases and controls.

**Results:** We identified a total of 158 RPVs in SCFA-sensing GPCR genes. The prevalence of RPV carriers was significantly higher in patients with HTN (OR=1.12, P=0.014) and MACEs (OR=1.18, P=0.009) than controls. In single GPCR gene analyses, RPVs in the FFAR2 gene (encoding GPR43) were associated with an increased risk of HTN (OR=1.23, P=0.005). RPVs in the HCAR2 gene (encoding GPR109A) were associated with a markedly increased risk of heart failure (OR=1.57, P=0.012).

**Conclusions:** These findings confirm and extend previous results from knockout animal models in a large population-based cohort, highlighting the potential of GPCRs as therapeutic targets for HTN and cardiovascular diseases in humans.

---

Gut microbial metabolites called short-chain fatty acids (SCFAs) confer protection against cardiovascular disease (CVD) and its primary risk factor, hypertension.^1^ Dietary fibers reach the large intestine undigested, where some are metabolized by fermentative microbes, releasing SCFAs as by-products. These metabolites lower blood pressure in humans and protect against cardiac hypertrophy and fibrosis in mice.^1, 2^ Proposed mechanisms include anti-inflammatory signaling activated by the binding of SCFAs to G-protein-coupled receptors (GPCRs, particularly *FFAR3*/GPR41, *FFAR2*/GPR43, and *HCAR2*/GPR109A), as suggested by previous mouse studies using knockout strains.^1, 3^ Here, we conducted a large-scale population-based study aimed at determining if rare pathogenic variants (RPVs) affecting SCFA-sensing GPCR genes increase the risk in humans of hypertension and major adverse cardiac events (MACE), including acute coronary syndrome, heart failure, and ischemic stroke.

We used the population healthcare, lifestyle, and whole-exome sequencing (WES) data from the UK Biobank, available for 393,649 participants of white-European genetic ancestry (**Figure A**). UK Biobank has approval from the Northwest Multi-Centre Research Ethics Committee and conforms to the Declaration of Helsinki principles. Informed written consent was given before the inclusion of subjects in the study. Rare (minor allele frequency <1%) variants in the GPCR genes *FFAR3, FFAR2* and *HCAR2* and their genotypes were extracted from quality- controlled WES data, based on genomic coordinates. We identified a subset of 158 RPVs with predicted high-impact functional consequences or missense-pathogenic likelihood scores (**Figure A**). Annotations were based on the Ensembl Variant Effect Predictor tool (http://www.ensembl.org/info/docs/tools/vep), as performed previously.^4^ Cases and controls for hypertension and MACE subtypes were identified based on diagnoses from hospital admissions (recorded as ICD-10 – International Classification of Disease 10^th^ version - codes), self-reported medical histories from health-related questionnaires, surgical records and death registries (**Figure A**). The characteristics of the study cohort are detailed in **Figure B**.

**Figure 1.**
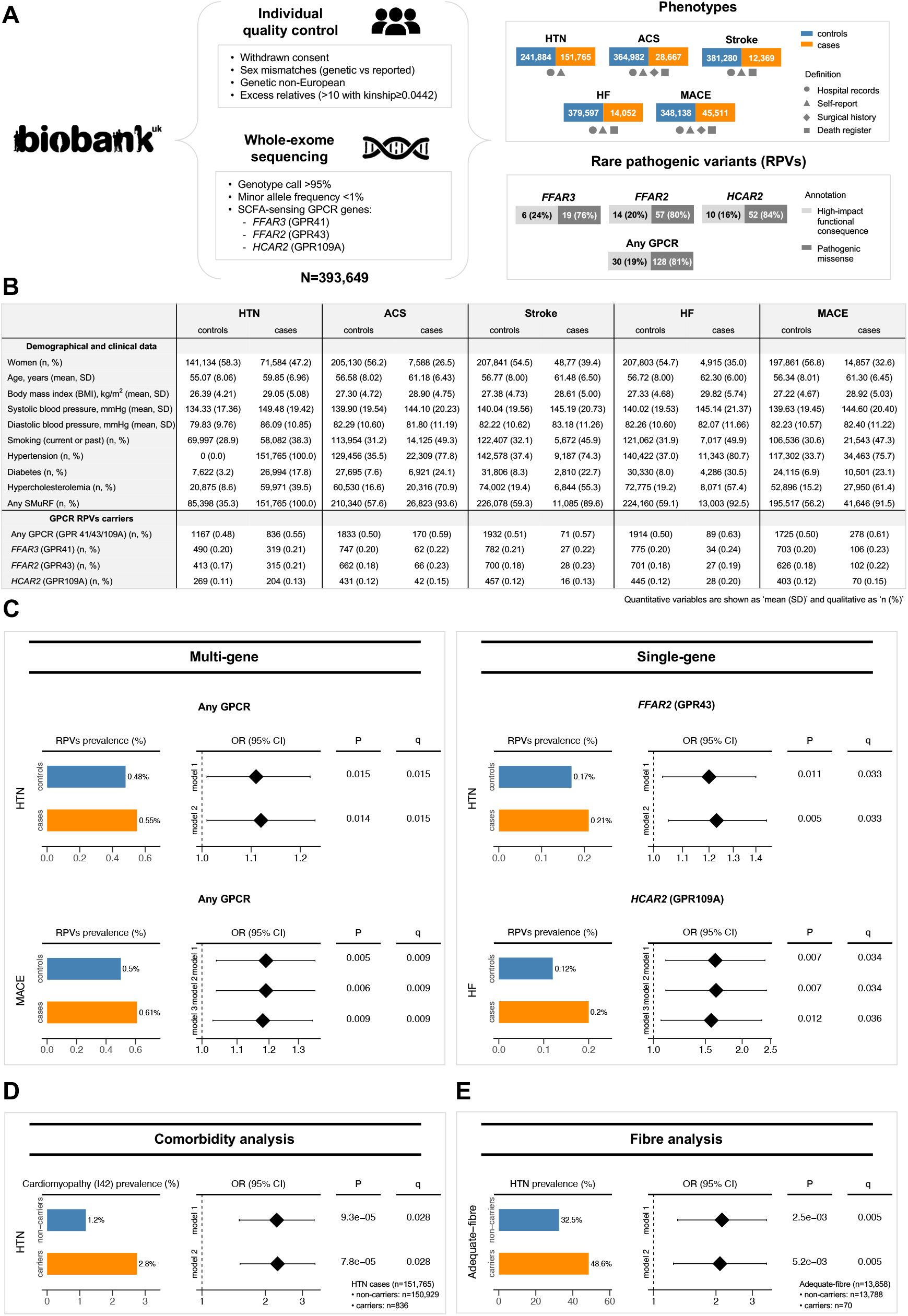
Rare pathogenic variants (RPVs) in short-chain fatty acid (SCFA)-sensing G- protein coupled receptors (GPCR) genes are associated with poor cardiovascular outcomes. **(A)** Study design flowchart, including phenotype and RPVs definitions. RPVs were identified based on two classifications: (1) *‘high-impact functional consequence’*, comprising stop gained/lost, frameshift, and splicing alterations based on Ensembl Variant Effect Predictor annotations; (2) *‘pathogenic missense variants’*, comprising missense variants with pathogenic likelihood scores >0.564 from AlphaMissense, >0.025 from Mendelian Clinically Applicable Pathogenicity, or >20 from Combined Annotation Dependent Depletion. **(B)** Study cohort demographic and clinical characteristics, stratified as cases and controls for each phenotype of interest, encompassing hypertension (HTN), major adverse cardiac events (MACE) and MACE subtypes (acute coronary syndrome (ACS), heart failure (HF), and ischemic stroke). **(C)** Significant associations between GPCR RPVs and phenotypes’ risk. **(D)** Increased cardiomyopathy prevalence in hypertensive cases carrying GPCR RPVs. **(E)** Increased hypertension prevalence in GPCR RPV carriers among adequate-fiber diet consumers. **Statistics:** Multiple comparisons were corrected by false discovery rate using the Benjamini- Hochberg method (threshold q<0.05). **Panels C-E:** Odds-ratio (OR) and P values are derived from multivariable logistic regression analyses, adjusted for: age and sex (*model 1*); age, sex, and BMI (*model 2*); age, sex, BMI, and standard modifiable cardiovascular risk factors (SMurFs), including smoking (current/past), hypertension, diabetes, and hypercholesterolemia (*model 3*; only applied for MACE and subtypes). Genetic ancestry (10-top genetic principal components) was also included as covariate when testing the association between GPCR RPVs and phenotypes’ risk (Panel C). All OR forest plots are in a log_10_ scale.

Multiple and single GPCR gene-based collapsing analyses were conducted to test whether there is a significant difference in the counts of individuals carrying at least one rare- pathogenic variant that disrupts gene function (carriers) between cases versus controls, using multivariable-adjusted regression models.^5^ Age- and sex-adjusted models, either excluding or including BMI as a covariate (*model 1* and *model 2*, respectively), revealed the prevalence of RPV carriers affecting any GPCR gene of interest was significantly higher in participants with hypertension and MACE compared to controls (**Figure C**). Given the occurrence of MACE is recognized to be associated with standard modifiable cardiovascular risk factors (SMuRFs) – including hypertension, diabetes, hypercholesterolemia, and smoking – we further adjusted MACE analyses for these additional covariates (*model 3*). SMuRFs-adjusted analyses confirmed a significantly increased risk of MACE among GPCR RPV carriers, independent of these additional risk factors (**Figure C**). Finally, when GPCR genes were tested individually, carrying RPVs in the *HCAR2* (GPR109A) gene was associated with a markedly increased risk of heart failure, while carrying RPVs in the *FFAR2* (GPR43) gene was associated with an increased risk of hypertension, with associations remaining significant after correction for multiple comparisons (accounting for 3 GPCR genes and distinct models) (**Figure C**).

Next, to identify other diseases and conditions that are more common among hypertension and MACE patients carrying GPCR RPVs (compared to non-carriers) and could contribute to this association, we performed a comorbidity analysis by retrieving hospital inpatient records for 1,259 ICD-10 codes (Chapters I–XIV) across 39,563,559 data-points (243 data-fields covering 162,813 cases for hypertension/MACE). After filtering for ICD-10 diseases with a prevalence greater than 1% and adjusting for multiple comparisons (297 ICD-10 diseases and two models), we found cardiomyopathy (ICD-10: I42) was more than twice as common in hypertensive patients carrying GPCR RPVs (**Figure D**). Consistently, cardiac hypertrophy and fibrosis have been observed in experimental mouse models that lack SCFA-sensing GPCR genes.^1, 3^

Since SCFAs are produced primarily by the gut microbiota as a response to dietary fiber intake, to further understand the complex relationship between fiber, gut-to-host communication, and cardiovascular health, we investigated whether carrying GPCR RPVs increases the risk of hypertension and MACE among individuals consuming an adequate-fiber diet. This would support the hypothesis that fiber’s beneficial cardiovascular effects depend upon GPCR signaling. To estimate daily fiber intake, we studied a subset of UK Biobank participants with dietary data, assessed by a 24-hour dietary recall of food intake on the previous day in five different instances (n=168,142 with dietary data available for at least one instance). Based on nutritional recommendations,^6^ an adequate-fiber diet was defined as an average intake of ≥25 g/day for women and ≥30 g/day for men, leaving 13,858 participants for subsequent analyses.

Consistent with our hypothesis, the prevalence of hypertension was significantly higher among GPCR RPVs carriers (**Figure E**), suggesting the cardiovascular benefits of fiber intake are diminished in the presence of genetic impairments in SCFA-related GPCR signaling. No significant associations were observed for MACE, likely due to the smaller sample size. Furthermore, in the inadequate-fiber consumers subgroup (n=154,284), hypertension prevalence was still higher in carriers versus non-carriers of RPVs (38.0% versus 34.9%), but it was no longer significant (p>0.05). In the absence of sufficient fiber, SCFA production is minimal and, thus, the benefit from SCFA signaling is likely blunted for both GPCR RPVs carriers and non- carriers.

Overall, our findings elucidate gene-by-environment interactions and show that lack of signaling via SCFA-sensing GPCRs, even when fiber intake is adequate, is associated with human hypertension and MACE in a large population-based cohort from the UK. This underscores the potential of targeting gut-to-host mechanisms through SCFA-sensing GPCRs as a therapeutic strategy for hypertension and CVD.

## Data Availability

This research has been conducted using the UK Biobank Resource under Application Number 86879. This work uses data provided by patients and collected by the NHS as part of their care and support.

## Acknowledgement

We would like to acknowledge Monash Bioinformatics Platform for access to the M3 servers, as well as the participants of the UK Biobank. This research has been conducted using the UK Biobank Resource under Application Number 86879.

