## Supplementary material for "Rare pathogenic variants in G-protein coupled receptor genes involved in gut-to-host communication are associated with cardiovascular disease risk": Online supplemental tables and figures

**Short title:** Gut-to-host receptors and cardiovascular disease risk

Leticia Camargo Tavares[1*]; Rikeish R. Muralitharan[1]; Matthew Snelson[1]; Francine Z. Marques[1,2*]

[1]Hypertension Research Laboratory, Victorian Heart Institute and Department of Pharmacology, Biomedical Discovery Institute, Faculty of Medicine, Nursing and Health Sciences, Monash University, Melbourne, Australia; [2]Heart Failure Research Laboratory, Baker Heart and Diabetes Institute, Melbourne, Australia.

***Corresponding authors**: Dr Leticia Camargo Tavares, PhD, Postdoctoral Research Fellow, Hypertension Research Laboratory, School of Biological Sciences, Faculty of Science, Monash University, Melbourne, Australia.; Prof Francine Marques, Hypertension Research Laboratory, Victorian Heart Institute, Level 2, Victorian Heart Hospital, 631 Blackburn Road Clayton, VIC 3168 Monash University, Melbourne, Australia, Phone: +61-03-9905 6958.

**Funding Information:** F.Z.M. is supported by a Senior Medical Research Fellowship from the Sylvia and Charles Viertel Charitable Foundation, a National Heart Foundation Future Leader Fellowship (105663), and National Health & Medical Research Council (NHMRC)

Emerging Leader Fellowship (GNT2017382). M.S. is supported by a National Heart Foundation Postdoctoral Fellowship (106698).

**Conflict of interest**

None.

**Data availability**

This research has been conducted using the UK Biobank Resource under Application Number 86879. This work uses data provided by patients and collected by the NHS as part of their care and support.

**Acknowledgement**

We would like to acknowledge Monash Bioinformatics Platform for access to the M3 servers, as well as the participants of the UK Biobank. This research has been conducted using the UK Biobank Resource under Application Number 86879.

**Author contributions**

LCT led and FZM supported study design and conception; LCT led the formal data analysis and visualization; LCT led and RRM, MS, and FZM supported data interpretation; LCT led and RRM, MS, and FZM supported drafting the manuscript; All authors approved the final version of the manuscript.

**Keywords**

Genomics; Rare variants; G-protein couple receptors; Short-chain fatty acids; Hypertension; Major adverse cardiac events.

### Supplementary Material

#### Supplementary Methods

UK Biobank study subjects…………………………………………………………………..4

Phenotypes definition………………………………………………………………………...4

Identification of rare pathogenic variants in SCFA-sensing GPCR genes…………………...5

Association analyses between RPVs carriership and phenotype risk………………………...6

Comorbidity analyses………………………………………………………………………...7

Fiber analyses………………………………………………………………………………...7

Table S8……………………………………………………………………………………...24

Table S9……………………………………………………………………………………...26

Table S10……………………………………………………………………………………..27

Table S11……………………………………………………………………………………..28

Table S12……………………………………………………………………………………..29

Table S13……………………………………………………………………………………..30

Table S14……………………………………………………………………………………..31

Table S15……………………………………………………………………………………..32

#### Supplementary Methods

##### UK Biobank study subjects

The UK Biobank (UKBB) is a population-based longitudinal cohort of >500,000 participants aged between 40 and 69 years, with available genotype, demographic and health-related data.[1] Health information encompasses electronic medical records (EMR) from hospital inpatient admissions, questionnaire data about general health and lifestyle, and self-report of medical diagnoses and medication usage. UKBB follows the principles of the Declaration of Helsinki and received ethical approval from the competent Research Ethics Committee (REC reference for UKBB is 11/NW/0382). The data was obtained under Application Number 86879 from UKBB.

Upon exclusion of individuals with withdrawn consent, sex mismatches (genetic versus reported), with excess relatives (>10 relatives with kinship≥0.0442) and of non-white-European genetic ancestry, a total of 393,649 participants with whole-exome sequencing (WES) data were included in downstream analyses (**Table S1**). Given the limited representation of individuals from other populations in the UKBB and to ensure meaningful statistical power, our analyses were restricted to individuals of white European genetic ancestry (identified through the data-field 22006).

##### Phenotype definitions

Participants were identified as patients (cases) with essential hypertension or major adverse cardiac events (MACE), including acute coronary syndrome (ACS), heart failure (HF), and ischemic stroke, as previously reported.[2] For each phenotype of interest, controls were defined as participants without the condition. UKBB data-fields utilized for hypertension and MACE phenotype definitions encompassed electronic medical records from hospital inpatient (either primary or secondary) episodes (data field 41270), self-reported medical diagnoses (data field 20002), death registers (data fields 40001 and 40002), and surgical records (data fields 41272 and 20004) related to revascularization procedures, including percutaneous coronary intervention or coronary artery bypass graft (detailed in **Table S2**). Specifically, essential hypertension was identified based on the hospital-inpatient records bearing the ICD-10 (International Classification of Diseases, 10th revision) code "I10" or through self-reported medical history (codes 1065 or 1072). The occurrence of MACE is recognized to be associated with SMuRFs, including hypertension, smoking, diabetes, and hypercholesterolemia.[3] The definitions of SMuRFs were determined based on inpatient diagnoses from medical records, self-reported medical histories, or the self-reported usage of insulin or cholesterol-lowering medication, as described in **Table S3**. Demographic and clinical characteristics of the study cohort are shown in **Table S4**.

**Identification of rare pathogenic variants (RPVs) in SCFA-sensing GPCR genes**

UKBB WES data were produced using the IDT xGen Exome Research Panel v1.0 on Illumina NovaSeq 6000 platform, which targets 39 megabases of the human genome (19,396 genes), with 20X coverage at 95% of sites on average.[4] Rare (minor allele frequency <1%) DNA variants in short-chain fatty acid (SCFA)-sensing G-protein coupled receptor (GPCR) genes were extracted from PLINK-formatted WES data (UKBB category 170) based on genomic coordinates encompassing *FFAR3*/GPR41, *FFAR2*/GPR43 and *HCAR2*/GPR109A gene regions according to the GRCh38 human genome build. Markers with call rates <95% were excluded. Variants' functional consequences were annotated with Ensembl Variant Effect Predictor (VEP, release 112, http://www.ensembl.org/info/docs/tools/vep).[5] Pathogenic variants were identified using two classification criteria: (1) '*high-impact functional consequence*', which includes stop gained/lost, frameshift, and splicing alterations based on Ensembl transcripts and Sequence Ontology terms (https://asia.ensembl.org/info/genome/variation/prediction/predicted_data.html)[5]; (2) '*pathogenic missense variants*', which includes missense variants with pathogenic likelihood scores >0.564 from AlphaMissense (http://alphamissense.hegelab.org)[6], >0.025 from M-CAP (Mendelian Clinically Applicable Pathogenicity, http://bejerano.stanford.edu/MCAP)[7], or >20 from CADD (Combined Annotation Dependent Depletion, http://cadd.gs.washington.edu/)[8] classifiers, as similarly performed previously.[9] In total, 158 variants were identified as rare pathogenic variants (RPVs) and included in downstream association analyses: 25 in *FFAR3/*GPR41 (24% high-impact, 76% pathogenic missense), 71 in *FFAR2/*GPR43 (20% high-impact, 80% pathogenic missense), and 62 in *HCAR2*/GPR109A (16% high impact, 84% pathogenic missense). VEP annotations are described in **Table S5**.

**Association analyses between RPVs carriership and phenotype risk**

Multiple and single GPCR gene-based collapsing analyses were conducted to test whether the presence of RPVs that disrupt gene function is associated with increased phenotype risk (based on one-tailed statistics), using multivariable-adjusted logistic regression models implemented in R (*glm* function). Variants meeting our pathogenicity criteria (outlined above) were binned together as equivalent to test whether their presence (or absence) is increased in cases relative to controls.[10] For these analyses, UKBB cases and controls were stratified into carriers of RPVs in any GPCR gene (for multiple gene-based analysis) or carriers of RPVs in individual GPCR genes (for single gene-based analysis) and compared with their respective non-carriers (**Table S6**). Age- and sex-adjusted models were conducted, either excluding or including BMI as a covariate (*model 1* and *model 2*, respectively). Given the occurrence of MACE is recognized to be associated with SMuRFs, we further adjusted MACE analyses for these additional covariates (*model 3*). Genetic ancestry captured by the 10-top genetic principal components (extracted as pre-computed PCs from data field 22020) was also included as covariate when testing the association between GPCR RPVs and phenotypes' risk, to account for population structure effects. Multiple comparisons were corrected by false discovery rate using the Benjamini-Hochberg method (threshold $q<0.05$) to control for type I error for each phenotype of interest. This accounted for the distinct models applied with different covariates (i.e., *model 1*, *model 2*, and *model 3*). For the single-gene analysis, we further corrected the significance threshold by the three SCFA-sensing GPCR genes tested individually (i.e., 3 genes * number of covariate models tested). Full results are shown in **Table S7** (multiple gene-based analysis) and **Table S8** (single gene-based analysis).

**Comorbidity analyses**

To identify other diseases and conditions that are more common among hypertension and MACE patients carrying GPCR RPVs (compared to non-carriers) and could contribute to RPVs-phenotype risk associations, we performed a comorbidity analysis by retrieving hospital inpatient records for 1,259 ICD-10 codes (Chapters I–XIV) across 39,563,559 data-points (243 data-fields covering 162,813 cases for hypertension/MACE). Only ICD-10 diseases with a prevalence greater than 1% were included, resulting in 297 ICD-10 diseases in the analyses. Age- and sex-adjusted models, either excluding or including BMI as a covariate (*model 1* and *model 2*, respectively), were applied to compare the prevalence of these ICD-10 comorbid diseases within cases of each phenotype of interest (n=162,813) stratified as GPCR RPVs carriers versus non-carriers. Demographic and clinical characteristics are detailed in **Table S9**. FDR adjustment for multiple comparisons was applied, accounting for 297 ICD-10 diseases and two models. Statistically significant results are shown in **Table S10**.

**Fiber analyses**

To further understand the complex relationship between fiber, gut-to-host communication, and cardiovascular health, we investigated whether carrying GPCR RPVs increases the risk of hypertension and MACE among individuals consuming an adequate-fiber diet. To estimate daily fiber intake, we studied a subset of UK Biobank participants with dietary data, assessed by a 24-hour dietary recall of food intake on the previous day in five different instances (n=168,142 with dietary data available for at least one instance). Based on nutritional recommendations,[11] an adequate-fiber diet was defined as an average intake of ≥25 g/day for women and ≥30 g/day for men. Demographic and clinical characteristics of adequate-fiber consumers (n=13,858) and inadequate-fiber consumers (n=154,284) are detailed in **Table S11, Table S13**, and **Table S15.** Age- and sex-adjusted models, either excluding or including BMI as a covariate (*model 1* and *model 2*, respectively), were conducted to compare the prevalence of hypertension between GPCR RPVs carriers versus non-carriers among adequate-fiber consumers and inadequate-fiber consumers (**Table S12** and **Table S14,** respectively).

### Supplementary Tables

**Table S1.** Individual quality control in the overall study cohort in UK Biobank.

| Filtering step | Samples removed | Samples left |
|---|---|---|
| Initial number | 0 | 502,411 |
| Remove individuals with withdrawn consent | -231 | 502,180 |
| WES data available | -32,598 | 469,582 |
| Sex mismatch check | -477 | 469,105 |
| Self-reported White individuals (British, Irish and other white) | -26,791 | 442,314 |
| Genetic white-European individuals (based on principal component analysis) | -48,626 | 393,688 |
| Remove individuals with excess (>10) putative third-degree relatives (kinship≥0.0442) | -39 | **393,649** |

**Table S2.** Phenotype definitions and their associated data-field and coding in UK Biobank.

| Phenotype definition | HTN | ACS | Stroke | HF |
|---|---|---|---|---|
| **Hospital inpatient diagnosis (ICD-10)** | 41270 in (I10.*) | 41270 in (I20.0, I21.*, I24.8, I24.9) | 41270 in (I63.*, I64.*) | 41270 in (I50.*) |
| **Self-reported medical history** | 20002 in (1065, 1072) | 20002 in (1075) | 20002 in (1081, 1583) | 20002 in (1076) |
| **Surgical code (OPCS-4)** | - | 41272 in (K40.*, K41.*, K42.*, K43.*, K44.*, K45.*, K46.*, K49.*, K50.*, K75.*) | - | - |
| **Self-reported surgical history** | - | 20004 in (1070, 1095) | - | - |
| **Death register** | - | 40001 in (I20.0, I21.*, I24.8, I24.9) or 40002 in (I20.0, I21.*, I24.8, I24.9) | 40001 in (I63.*, I64.*) or 40002 in (I63.*, I64.*) | 40001 in (I50.*) or 40002 in (I50.*) |

**Legend:** Phenotype definitions associated data-field and coding in UK Biobank are shown as 'data-field number in (coding number)'

**ACS:** Acute coronary syndrome; **HTN:** Essential hypertension; **HF:** Heart failure; **ICD:** International Classification of Diseases; **OPCS:** Operating Procedure Codes Supplement.

**Table S3.** Standard modifiable cardiovascular risk factors (SMuRFs) definitions and their associated data-field and coding in UK Biobank.

| Phenotype definition | Smoking (current or past) | HTN | Diabetes | Hypercholesterolaemia |
|---|---|---|---|---|
| **Hospital inpatient diagnosis (ICD-10)** | - | 41270 in (I10.*) | 41270 in (E10.*, E11.*, E12.*, E13.*, E14.*) | 41270 in (E78.0) |
| **Self-reported medical history** | - | 20002 in (1065, 1072) | 20002 in (1220, 1221, 1222, 1223) | - |
| **Self-reported medication** | - | - | 6177 in (3) | 6177 in (1) |
| **Smoking lifestyle** | 1239 in (1) or 1249 in (1) | - | - | - |

**Legend**: Phenotype definitions associated data-field and coding in UK Biobank are shown as ‘data-field number in (coding number)’

**HTN:** Essential hypertension; **ICD:** International Classification of Diseases.

**Table S4.** Study cohort demographic and clinical characteristics (n=393,649).

| | HTN | | ACS | | Stroke | | HF | | MACE | |
|---|---|---|---|---|---|---|---|---|---|---|
| | controls | cases | controls | cases | controls | cases | controls | cases | controls | cases |
| N of participants | 241,884 | 151,765 | 364,982 | 28,667 | 381,280 | 12,369 | 379,597 | 14,052 | 348,138 | 45,511 |
| Women (n, %) | 141,134 (58.3) | 71,584 (47.2) | 205,130 (56.2) | 7,588 (26.5) | 207,841 (54.5) | 48,77 (39.4) | 207,803 (54.7) | 4,915 (35.0) | 197,861 (56.8) | 14,857 (32.6) |
| Age, years (mean, SD) | 55.07 (8.06) | 59.85 (6.96) | 56.58 (8.02) | 61.18 (6.43) | 56.77 (8.00) | 61.48 (6.50) | 56.72 (8.00) | 62.30 (6.00) | 56.34 (8.01) | 61.30 (6.45) |
| BMI, kg/m$^2$ (mean, SD) | 26.39 (4.21) | 29.05 (5.08) | 27.30 (4.72) | 28.90 (4.75) | 27.38 (4.73) | 28.61 (5.00) | 27.33 (4.68) | 29.82 (5.74) | 27.22 (4.67) | 28.92 (5.03) |
| SBP, mmHG (mean, SD) | 134.33 (17.36) | 149.48 (19.42) | 139.90 (19.54) | 144.10 (20.23) | 140.04 (19.56) | 145.19 (20.73) | 140.02 (19.53) | 145.14 (21.37) | 139.63 (19.45) | 144.60 (20.40) |
| DBP, mmHG (mean, SD) | 79.83 (9.76) | 86.09 (10.85) | 82.29 (10.60) | 81.80 (11.19) | 82.22 (10.62) | 83.18 (11.26) | 82.26 (10.60) | 82.07 (11.66) | 82.23 (10.57) | 82.40 (11.22) |
| Smoking (current or past) | 69,997 (28.9) | 58,082 (38.3) | 113,954 (31.2) | 14,125 (49.3) | 122,407 (32.1) | 5,672 (45.9) | 121,062 (31.9) | 7,017 (49.9) | 106,536 (30.6) | 21,543 (47.3) |
| HTN (n, %) | 0 (0.0) | 151,765 (100.0) | 129,456 (35.5) | 22,309 (77.8) | 142,578 (37.4) | 9,187 (74.3) | 140,422 (37.0) | 11,343 (80.7) | 117,302 (33.7) | 34,463 (75.7) |
| Diabetes (n, %) | 7,622 (3.2) | 26,994 (17.8) | 27,695 (7.6) | 6,921 (24.1) | 31,806 (8.3) | 2,810 (22.7) | 30,330 (8.0) | 4,286 (30.5) | 24,115 (6.9) | 10,501 (23.1) |
| Hypercholesterolemia (n, %) | 20,875 (8.6) | 59,971 (39.5) | 60,530 (16.6) | 20,316 (70.9) | 74,002 (19.4) | 6,844 (55.3) | 72,775 (19.2) | 8,071 (57.4) | 52,896 (15.2) | 27,950 (61.4) |
| Any SMuRF (n, %) | 85,398 (35.3) | 151,765 (100.0) | 210,340 (57.6) | 26,823 (93.6) | 226,078 (59.3) | 11,085 (89.6) | 224,160 (59.1) | 13,003 (92.5) | 195,517 (56.2) | 41,646 (91.5) |

**Legend: ACS:** Acute coronary syndrome; **BMI:** Body mass index; **HTN:** Essential hypertension; **HF:** Heart failure; **MACE:** Major adverse cardiac events; **SD:** Standard deviation; **SBP:** Systolic blood pressure; **DBP:** Diastolic blood pressure; **SMurFs:** Standard modifiable cardiovascular risk factor, including smoking, HTN, diabetes, and hypercholesterolemia.

**Table S5.** GPCR RPVs annotations obtained with the Ensembl Variant Effect Predictor tool.

| GPCR gene | Variant (CHR:BP:OA:EA) | rsID | MAF | Pathogenic group | Functional consequence | Protein position | Amino acid change | AlphaMissense score | M-CAP score | CADD phred score |
|---|---|---|---|---|---|---|---|---|---|---|
| *HCAR2* | 12:122702192:T:G | | 1.270E-06 | High-impact | stop_lost | 364 | */Y | | | 6.18 |
| *HCAR2* | 12:122702194:A:C | | 1.270E-06 | High-impact | stop_lost | 364 | */E | | | 4.83 |
| *HCAR2* | 12:122702381:A:C | | 2.540E-06 | Pathogenic missense | missense_variant | 301 | F/L | 0.9724 | 0.0441 | 15.18 |
| *HCAR2* | 12:122702413:G:T | | 1.270E-06 | Pathogenic missense | missense_variant | 291 | P/T | 0.9632 | 0.5408 | 25.1 |
| *HCAR2* | 12:122702516:C:G | | 5.081E-06 | Pathogenic missense | missense_variant | 256 | W/C | 0.6014 | 0.0147 | 22.5 |
| *HCAR2* | 12:122702516:C:T | | 3.811E-06 | High-impact | stop_gained | 256 | W/* | | | 35 |
| *HCAR2* | 12:122702533:G:A | rs776840309 | 5.081E-06 | Pathogenic missense | missense_variant | 251 | R/W | 0.6285 | 0.0664 | 23.9 |
| *HCAR2* | 12:122702542:C:G | rs775536264 | 2.540E-06 | Pathogenic missense | missense_variant | 248 | V/L | 0.7207 | 0.0102 | 22.1 |
| *HCAR2* | 12:122702543:G:T | rs201091527 | 1.816E-04 | Pathogenic missense | missense_variant | 247 | S/R | 0.996 | 0.1751 | 17.47 |
| *HCAR2* | 12:122702544:C:T | rs1877107348 | 5.081E-06 | Pathogenic missense | missense_variant | 247 | S/N | 0.9134 | 0.062 | 24.6 |
| *HCAR2* | 12:122702554:A:G | rs202134216 | 8.764E-05 | Pathogenic missense | missense_variant | 244 | F/L | 0.9982 | 0.2401 | 26.7 |
| *HCAR2* | 12:122702609:C:A | rs201310632 | 1.232E-04 | Pathogenic missense | missense_variant | 225 | K/N | 0.7504 | 0.018 | 22.8 |
| *HCAR2* | 12:122702629:G:A | rs200587011 | 7.621E-06 | High-impact | stop_gained | 219 | Q/* | | | 35 |
| *HCAR2* | 12:122702659:C:G | | 2.540E-06 | Pathogenic missense | missense_variant | 209 | A/P | 0.6986 | 0.0439 | 16.79 |
| *HCAR2* | 12:122702665:A:G | | 1.270E-06 | Pathogenic missense | missense_variant | 207 | C/R | 0.9928 | 0.0552 | 26.3 |
| *HCAR2* | 12:122702697:T:A | | 1.270E-06 | Pathogenic missense | missense_variant | 196 | E/V | 0.6775 | 0.1527 | 25.2 |
| *HCAR2* | 12:122702700:A:G | rs200492929 | 5.081E-06 | Pathogenic missense | missense_variant | 195 | L/P | 0.8108 | 0.0432 | 26.3 |
| *HCAR2* | 12:122702716:C:T | rs200721962 | 8.891E-06 | Pathogenic missense | missense_variant | 190 | E/K | 0.5779 | 0.0133 | 17.62 |
| *HCAR2* | 12:122702720:C:G | rs1341264802 | 2.540E-06 | Pathogenic missense | missense_variant | 188 | W/C | 0.9514 | 0.0845 | 24.5 |
| *HCAR2* | 12:122702736:C:G | rs201831072 | 7.621E-06 | Pathogenic missense | missense_variant | 183 | C/S | 0.9159 | 0.0192 | 20.3 |
| *HCAR2* | 12:122702754:C:T | rs1194187180 | 2.540E-06 | Pathogenic missense | missense_variant | 177 | C/Y | 0.9819 | 0.2813 | 22.8 |
| *HCAR2* | 12:122702754:CA:C | | 1.270E-06 | High-impact | frameshift_variant | 177 | C/X | | | |
| *HCAR2* | 12:122702814:C:T | | 1.270E-06 | Pathogenic missense | missense_variant | 157 | G/D | 0.8723 | 0.0222 | 18.62 |

| | | | | | | | | | | |
|---|---|---|---|---|---|---|---|---|---|---|
| *HCAR2* | 12:122702825:GC:G | | 1.270E-06 | High-impact | frameshift_variant | 153 | G/X | | | |
| *HCAR2* | 12:122702841:G:A | | 1.270E-06 | Pathogenic missense | missense_variant | 148 | S/F | 0.9559 | 0.045 | 26.2 |
| *HCAR2* | 12:122702899:C:G | | 7.621E-06 | Pathogenic missense | missense_variant | 129 | V/L | 0.9334 | 0.1078 | 23 |
| *HCAR2* | 12:122702902:G:A | rs201756503 | 6.351E-06 | Pathogenic missense | missense_variant | 128 | R/W | 0.6971 | 0.089 | 23.2 |
| *HCAR2* | 12:122702919:G:A | rs200705418 | 1.143E-05 | Pathogenic missense | missense_variant | 122 | A/V | 0.7908 | 0.1984 | 24.7 |
| *HCAR2* | 12:122702928:G:C | | 1.143E-05 | Pathogenic missense | missense_variant | 119 | T/R | 0.9815 | 0.2598 | 24.2 |
| *HCAR2* | 12:122702933:G:T | rs199839290 | 1.270E-05 | Pathogenic missense | missense_variant | 117 | F/L | 0.9802 | 0.0119 | 26.2 |
| *HCAR2* | 12:122702940:A:T | | 1.270E-06 | Pathogenic missense | missense_variant | 115 | I/N | 0.9087 | 0.2487 | 25.2 |
| *HCAR2* | 12:122702947:C:T | | 1.270E-06 | Pathogenic missense | missense_variant | 113 | G/S | 0.5859 | 0.0425 | 24.4 |
| *HCAR2* | 12:122702953:G:A | rs753670084 | 3.811E-06 | Pathogenic missense | missense_variant | 111 | R/C | 0.9087 | 0.1627 | 26.7 |
| *HCAR2* | 12:122702963:C:A | rs752639607 | 6.351E-06 | Pathogenic missense | missense_variant | 107 | L/F | 0.6978 | 0.0438 | 22.1 |
| *HCAR2* | 12:122702964:A:C | rs1234650625 | 1.270E-06 | Pathogenic missense | missense_variant | 107 | L/W | 0.8949 | 0.104 | 25.9 |
| *HCAR2* | 12:122702982:C:G | | 1.270E-06 | Pathogenic missense | missense_variant | 101 | R/P | 0.9474 | 0.1481 | 24.2 |
| *HCAR2* | 12:122702983:G:A | rs200794525 | 1.016E-05 | Pathogenic missense | missense_variant | 101 | R/W | 0.6352 | 0.1087 | 24.1 |
| *HCAR2* | 12:122702997:C:T | | 1.270E-06 | Pathogenic missense | missense_variant | 96 | G/E | 0.671 | 0.0267 | 23.3 |
| *HCAR2* | 12:122702999:A:C | | 1.270E-06 | Pathogenic missense | missense_variant | 95 | F/L | 0.9333 | 0.0251 | 20.8 |
| *HCAR2* | 12:122703006:C:T | rs770293961 | 1.270E-06 | High-impact | stop_gained | 93 | W/* | | | 36 |
| *HCAR2* | 12:122703011:C:T | | 1.270E-06 | High-impact | stop_gained | 91 | W/* | | | 27.8 |
| *HCAR2* | 12:122703015:C:G | rs200252776 | 8.891E-06 | Pathogenic missense | missense_variant | 90 | R/P | 0.6079 | 0.0158 | 10.97 |
| *HCAR2* | 12:122703026:G:T | | 7.621E-06 | Pathogenic missense | missense_variant | 86 | N/K | 0.9259 | 0.041 | 13.16 |
| *HCAR2* | 12:122703028:T:C | | 3.811E-06 | Pathogenic missense | missense_variant | 86 | N/D | 0.6015 | 0.058 | 18.32 |
| *HCAR2* | 12:122703033:A:T | rs1396108170 | 3.811E-06 | Pathogenic missense | missense_variant | 84 | M/K | 0.7304 | 0.0111 | 7.774 |
| *HCAR2* | 12:122703057:A:G | COSV105903496 | 1.270E-06 | Pathogenic missense | missense_variant | 76 | L/P | 0.9889 | 0.1396 | 25.6 |
| *HCAR2* | 12:122703070:C:T | COSV61024552 | 1.270E-06 | Pathogenic missense | missense_variant | 72 | A/T | 0.623 | 0.1256 | 25.2 |
| *HCAR2* | 12:122703080:G:C | | 1.270E-06 | Pathogenic missense | missense_variant | 68 | N/K | 0.9954 | 0.0167 | 25.5 |
| *HCAR2* | 12:122703096:C:G | | 1.270E-06 | Pathogenic missense | missense_variant | 63 | R/P | 0.8998 | 0.0213 | 23 |
| *HCAR2* | 12:122703100:T:G | rs530458347 | 2.540E-06 | Pathogenic missense | missense_variant | 62 | S/R | 0.9809 | 0.0634 | 25.9 |

| | | | | | | | | | | |
|---|---|---|---|---|---|---|---|---|---|---|
| *HCAR2* | 12:122703126:C:T | | 1.270E-06 | Pathogenic missense | missense_variant | 53 | C/Y | 0.6438 | 0.0114 | 25.1 |
| *HCAR2* | 12:122703142:C:T | | 1.270E-06 | Pathogenic missense | missense_variant | 48 | A/T | 0.8578 | 0.0417 | 25.5 |
| *HCAR2* | 12:122703147:C:T | | 1.270E-06 | Pathogenic missense | missense_variant | 46 | G/D | 0.9864 | 0.016 | 24.1 |
| *HCAR2* | 12:122703150:T:A | rs763999055 | 1.270E-06 | Pathogenic missense | missense_variant | 45 | N/I | 0.9688 | 0.484 | 25.7 |
| *HCAR2* | 12:122703154:C:A | rs767733941 | 1.270E-06 | Pathogenic missense | missense_variant | 44 | G/C | 0.8324 | 0.046 | 24.1 |
| *HCAR2* | 12:122703163:C:G | rs1348029018 | 1.270E-06 | Pathogenic missense | missense_variant | 41 | G/R | 0.9955 | 0.1124 | 23.9 |
| *HCAR2* | 12:122703163:C:T | rs1348029018 | 1.270E-06 | Pathogenic missense | missense_variant | 41 | G/R | 0.9955 | 0.1195 | 24 |
| *HCAR2* | 12:122703173:C:CT | | 1.270E-06 | High-impact | frameshift_variant | 37 | E/EX | | | |
| *HCAR2* | 12:122703177:A:G | rs780534579 | 6.351E-06 | Pathogenic missense | missense_variant | 36 | L/P | 0.9096 | 0.0848 | 23.3 |
| *HCAR2* | 12:122703181:C:T | | 1.270E-06 | Pathogenic missense | missense_variant | 35 | G/R | 0.9372 | 0.0403 | 23.4 |
| *HCAR2* | 12:122703192:G:A | rs141014410 | 7.621E-06 | Pathogenic missense | missense_variant | 31 | P/L | 0.6174 | 0.0439 | 24 |
| *HCAR2* | 12:122703220:G:A | rs764643885 | 6.351E-06 | High-impact | stop_gained | 22 | R/* | | | 35 |
| *FFAR3* | 19:35359043:C:A | rs372231935 | 1.270E-06 | Pathogenic missense | missense_variant | 51 | D/E | 0.6816 | 0.0075 | 17.83 |
| *FFAR3* | 19:35359071:G:A | rs139036566 | 2.159E-05 | Pathogenic missense | missense_variant | 61 | D/N | 0.6195 | 0.0353 | 25.2 |
| *FFAR3* | 19:35359105:T:C | rs202122950 | 8.358E-04 | Pathogenic missense | missense_variant | 72 | M/T | 0.6665 | 0.0196 | 24 |
| *FFAR3* | 19:35359133:G:A | rs759261993 | 1.524E-05 | High-impact | stop_gained | 81 | W/* | | | 35 |
| *FFAR3* | 19:35359183:C:T | rs1332664480 | 3.811E-06 | Pathogenic missense | missense_variant | 98 | T/I | 0.6887 | 0.0313 | 22.3 |
| *FFAR3* | 19:35359298:T:G | | 6.351E-06 | Pathogenic missense | missense_variant | 136 | S/R | 0.9858 | 0.0648 | 8.091 |
| *FFAR3* | 19:35359330:G:A | rs752926781 | 1.016E-05 | Pathogenic missense | missense_variant | 147 | C/Y | 0.7885 | 0.0143 | 25.9 |
| *FFAR3* | 19:35359396:G:A | rs370522843 | 3.811E-06 | Pathogenic missense | missense_variant | 169 | C/Y | 0.872 | 0.0634 | 24.9 |
| *FFAR3* | 19:35359419:C:T | rs546623697 | 7.621E-06 | High-impact | stop_gained | 177 | Q/* | | | 34 |
| *FFAR3* | 19:35359505:C:A | rs770864257 | 4.573E-05 | Pathogenic missense | missense_variant | 205 | S/R | 0.6682 | 0.015 | 22.6 |
| *FFAR3* | 19:35359529:AG:A | rs759936957 | 3.811E-06 | High-impact | frameshift_variant | 214 | G/X | | | |
| *FFAR3* | 19:35359579:C:A | | 2.540E-06 | Pathogenic missense | missense_variant | 230 | T/K | 0.8242 | 0.0146 | 24.1 |
| *FFAR3* | 19:35359592:C:G | | 6.351E-06 | Pathogenic missense | missense_variant | 234 | F/L | 0.9407 | 0.0157 | 23.4 |
| *FFAR3* | 19:35359600:G:C | | 2.540E-06 | Pathogenic missense | missense_variant | 237 | C/S | 0.6139 | 0.0147 | 25.5 |
| *FFAR3* | 19:35359604:T:G | | 3.811E-06 | Pathogenic missense | missense_variant | 238 | F/L | 0.9499 | 0.0426 | 22.4 |

| | | | | | | | | | | |
|---|---|---|---|---|---|---|---|---|---|---|
| *FFAR3* | 19:35359605:G:A | | 2.540E-06 | Pathogenic missense | missense_variant | 239 | G/R | 0.9361 | 0.0465 | 26.1 |
| *FFAR3* | 19:35359621:C:T | rs561011815 | 8.891E-06 | Pathogenic missense | missense_variant | 244 | S/F | 0.8629 | 0.0604 | 24 |
| *FFAR3* | 19:35359625:T:G | | 1.270E-06 | Pathogenic missense | missense_variant | 245 | H/Q | 0.6576 | 0.0094 | 0.316 |
| *FFAR3* | 19:35359690:T:C | rs2066984319 | 6.351E-06 | Pathogenic missense | missense_variant | 267 | L/P | 0.7516 | 0.042 | 26.6 |
| *FFAR3* | 19:35359696:C:T | rs767908892 | 2.540E-06 | Pathogenic missense | missense_variant | 269 | S/F | 0.7431 | 0.0588 | 24.7 |
| *FFAR3* | 19:35359704:G:A | rs369542605 | 8.891E-06 | Pathogenic missense | missense_variant | 272 | D/N | 0.7315 | 0.0213 | 26.2 |
| *FFAR3* | 19:35359749:T:C | rs1261433323 | 5.081E-06 | Pathogenic missense | missense_variant | 287 | F/L | 0.572 | 0.0036 | 14.8 |
| *FFAR3* | 19:35359793:G:A | | 2.540E-06 | High-impact | stop_gained | 301 | W/* | | | 33 |
| *FFAR3* | 19:35359854:C:T | rs776486107 | 1.778E-05 | High-impact | stop_gained | 322 | R/* | | | 27.9 |
| *FFAR3* | 19:35359885:C:CA | | 2.540E-06 | High-impact | frameshift_variant | 332 | S/SX | | | |
| *FFAR2* | 19:35449746:T:C | | 1.270E-06 | Pathogenic missense | missense_variant | 11 | L/P | 0.6625 | 0.051 | 24.2 |
| *FFAR2* | 19:35449761:T:A | | 1.270E-06 | Pathogenic missense | missense_variant | 16 | I/N | 0.5785 | 0.0438 | 25.6 |
| *FFAR2* | 19:35449766:T:C | | 1.270E-06 | Pathogenic missense | missense_variant | 18 | F/L | 0.7472 | 0.0156 | 23.1 |
| *FFAR2* | 19:35449776:G:A | rs2067367647 | 1.270E-06 | Pathogenic missense | missense_variant | 21 | G/D | 0.9685 | 0.0433 | 28.3 |
| *FFAR2* | 19:35449779:T:TCCCTGCCAACCTCCTGGCCC | | 3.811E-06 | High-impact | frameshift_variant | 22 | L/LPANLLAX | | | |
| *FFAR2* | 19:35449789:C:A | rs533923897 | 7.621E-06 | Pathogenic missense | missense_variant | 25 | N/K | 0.9711 | 0.2488 | 23.2 |
| *FFAR2* | 19:35449794:TG:T | | 3.811E-06 | High-impact | frameshift_variant | 27 | L/X | | | |
| *FFAR2* | 19:35449796:G:A | | 1.270E-06 | Pathogenic missense | missense_variant | 28 | A/T | 0.6035 | 0.0363 | 27.7 |
| *FFAR2* | 19:35449851:T:C | rs771538131 | 1.270E-06 | Pathogenic missense | missense_variant | 46 | I/T | 0.7425 | 0.0603 | 26.2 |
| *FFAR2* | 19:35449854:T:G | | 1.270E-06 | Pathogenic missense | missense_variant | 47 | L/R | 0.8835 | 0.1254 | 26.7 |
| *FFAR2* | 19:35449863:G:T | | 1.270E-06 | Pathogenic missense | missense_variant | 50 | S/I | 0.8803 | 0.0597 | 23.3 |
| *FFAR2* | 19:35449869:C:A | rs1253813043 | 2.540E-06 | Pathogenic missense | missense_variant | 52 | T/K | 0.9398 | 0.0594 | 25.3 |
| *FFAR2* | 19:35449872:T:C | | 1.270E-06 | Pathogenic missense | missense_variant | 53 | L/P | 0.896 | 0.0707 | 27.4 |
| *FFAR2* | 19:35449877:G:A | rs747543025 | 3.938E-05 | Pathogenic missense | missense_variant | 55 | D/N | 0.8827 | 0.1327 | 31 |
| *FFAR2* | 19:35449916:G:A | rs765581638 | 5.081E-06 | Pathogenic missense | missense_variant | 68 | E/K | 0.8934 | 0.0574 | 24.9 |
| *FFAR2* | 19:35449916:G:C | | 1.270E-06 | Pathogenic missense | missense_variant | 68 | E/Q | 0.5731 | 0.0426 | 24.1 |
| *FFAR2* | 19:35449918:G:C | | 1.270E-06 | Pathogenic missense | missense_variant | 68 | E/D | 0.8989 | 0.0543 | 22.5 |

| | | | | | | | | | | |
|---|---|---|---|---|---|---|---|---|---|---|
| *FFAR2* | 19:35449942:CCTGCCCAAGGTCGT:C | rs753777712 | 6.351E-06 | High-impact | frameshift_variant | 77-81 | LPKVV/X | | | 25.4 |
| *FFAR2* | 19:35449967:AC:A | | 2.540E-06 | High-impact | frameshift_variant | 85 | T/X | | | |
| *FFAR2* | 19:35449971:GT:G | rs1433117229 | 6.478E-05 | High-impact | frameshift_variant | 86 | S/X | | | |
| *FFAR2* | 19:35449974:T:C | | 2.540E-06 | Pathogenic missense | missense_variant | 87 | F/S | 0.7633 | 0.048 | 25.6 |
| *FFAR2* | 19:35449976:G:C | rs200320229 | 1.080E-04 | Pathogenic missense | missense_variant | 88 | G/R | 0.9719 | 0.018 | 23.7 |
| *FFAR2* | 19:35449979:T:G | rs1338150311 | 2.540E-06 | Pathogenic missense | missense_variant | 89 | F/V | 0.8007 | 0.0578 | 26.3 |
| *FFAR2* | 19:35449989:G:A | rs372925283 | 1.778E-05 | Pathogenic missense | missense_variant | 92 | S/N | 0.7506 | 0.0085 | 23.4 |
| *FFAR2* | 19:35450016:C:A | rs763737717 | 5.716E-05 | Pathogenic missense | missense_variant | 101 | A/E | 0.9186 | 0.0855 | 24.9 |
| *FFAR2* | 19:35450030:G:A | rs140779660 | 3.811E-06 | Pathogenic missense | missense_variant | 106 | E/K | 0.8892 | 0.0717 | 27.7 |
| *FFAR2* | 19:35450030:G:C | rs140779660 | 1.270E-05 | Pathogenic missense | missense_variant | 106 | E/Q | 0.6105 | 0.0315 | 26.1 |
| *FFAR2* | 19:35450033:C:CT | | 1.270E-06 | High-impact | frameshift_variant | 107 | R/LX | | | |
| *FFAR2* | 19:35450033:C:T | rs150111307 | 1.143E-05 | Pathogenic missense | missense_variant | 107 | R/C | 0.7818 | 0.4213 | 29 |
| *FFAR2* | 19:35450034:G:A | rs758560714 | 3.811E-06 | Pathogenic missense | missense_variant | 107 | R/H | 0.6966 | 0.2659 | 30 |
| *FFAR2* | 19:35450034:G:T | rs758560714 | 1.270E-06 | Pathogenic missense | missense_variant | 107 | R/L | 0.836 | 0.3919 | 31 |
| *FFAR2* | 19:35450045:G:T | | 1.270E-06 | Pathogenic missense | missense_variant | 111 | V/L | 0.7195 | 0.0495 | 25.4 |
| *FFAR2* | 19:35450052:T:C | rs1044871339 | 5.081E-06 | Pathogenic missense | missense_variant | 113 | F/S | 0.8855 | 0.032 | 24.5 |
| *FFAR2* | 19:35450116:G:T | rs1029056131 | 2.540E-06 | Pathogenic missense | missense_variant | 134 | W/C | 0.8036 | 0.3124 | 24.1 |
| *FFAR2* | 19:35450123:T:C | rs780672443 | 2.540E-06 | Pathogenic missense | missense_variant | 137 | S/P | 0.7827 | 0.0424 | 24.8 |
| *FFAR2* | 19:35450126:T:C | rs1298865522 | 7.621E-06 | Pathogenic missense | missense_variant | 138 | F/L | 0.6622 | 0.0066 | 21.8 |
| *FFAR2* | 19:35450130:G:A | | 2.540E-06 | Pathogenic missense | missense_variant | 139 | G/D | 0.9337 | 0.0128 | 24.4 |
| *FFAR2* | 19:35450137:C:A | rs769108749 | 2.540E-06 | High-impact | stop_gained | 141 | C/* | | | 35 |
| *FFAR2* | 19:35450144:G:A | rs1251799501 | 2.540E-06 | Pathogenic missense | missense_variant | 144 | V/M | 0.6047 | 0.0163 | 24.2 |
| *FFAR2* | 19:35450148:T:A | rs779367504 | 3.811E-06 | Pathogenic missense | missense_variant | 145 | I/N | 0.6214 | 0.0618 | 23.2 |
| *FFAR2* | 19:35450177:C:T | rs1330779175 | 6.351E-06 | High-impact | stop_gained | 155 | Q/* | | | 33 |
| *FFAR2* | 19:35450204:T:G | rs2067371834 | 1.270E-06 | Pathogenic missense | missense_variant | 164 | C/G | 0.942 | 0.0723 | 24 |
| *FFAR2* | 19:35450228:C:T | | 3.811E-06 | High-impact | stop_gained | 172 | Q/* | | | 36 |
| *FFAR2* | 19:35450252:C:T | | 1.270E-06 | Pathogenic missense | missense_variant | 180 | R/W | 0.7406 | 0.0725 | 23.7 |

| | | | | | | | | | | |
|---|---|---|---|---|---|---|---|---|---|---|
| *FFAR2* | 19:35450253:G:A | rs766489704 | 3.811E-06 | Pathogenic missense | missense_variant | 180 | R/Q | 0.652 | 0.0356 | 27.1 |
| *FFAR2* | 19:35450271:T:A | rs765896643 | 3.811E-06 | Pathogenic missense | missense_variant | 186 | V/E | 0.8507 | 0.0483 | 27 |
| *FFAR2* | 19:35450298:C:A | | 1.270E-06 | Pathogenic missense | missense_variant | 195 | T/N | 0.6187 | 0.0157 | 23.4 |
| *FFAR2* | 19:35450309:T:G | | 1.270E-06 | Pathogenic missense | missense_variant | 199 | Y/D | 0.7809 | 0.0999 | 28.3 |
| *FFAR2* | 19:35450316:G:GT | | 1.270E-06 | High-impact | frameshift_variant | 201 | R/RX | | | |
| *FFAR2* | 19:35450328:T:A | | 1.270E-06 | Pathogenic missense | missense_variant | 205 | I/N | 0.626 | 0.06 | 25.3 |
| *FFAR2* | 19:35450328:T:C | rs201319651 | 2.921E-05 | Pathogenic missense | missense_variant | 205 | I/T | 0.7213 | 0.015 | 24.4 |
| *FFAR2* | 19:35450369:C:G | rs1422005865 | 2.540E-06 | Pathogenic missense | missense_variant | 219 | R/G | 0.6559 | 0.0475 | 22.7 |
| *FFAR2* | 19:35450400:T:C | | 1.270E-06 | Pathogenic missense | missense_variant | 229 | L/P | 0.8908 | 0.0666 | 25.1 |
| *FFAR2* | 19:35450409:T:C | rs2067373826 | 8.891E-06 | Pathogenic missense | missense_variant | 232 | L/P | 0.9145 | 0.1695 | 25 |
| *FFAR2* | 19:35450420:G:A | rs143616979 | 3.899E-04 | Pathogenic missense | missense_variant | 236 | G/R | 0.9839 | 0.0735 | 23.2 |
| *FFAR2* | 19:35450435:T:C | rs2067374238 | 1.270E-06 | Pathogenic missense | missense_variant | 241 | S/P | 0.9808 | 0.0852 | 26.7 |
| *FFAR2* | 19:35450450:T:C | rs1419317668 | 1.143E-05 | Pathogenic missense | missense_variant | 246 | Y/H | 0.731 | 0.0107 | 22.8 |
| *FFAR2* | 19:35450468:C:T | | 5.081E-06 | Pathogenic missense | missense_variant | 252 | P/S | 0.7014 | 0.0555 | 23 |
| *FFAR2* | 19:35450476:G:T | | 1.270E-06 | Pathogenic missense | missense_variant | 254 | W/C | 0.9931 | 0.0883 | 26.5 |
| *FFAR2* | 19:35450477:C:T | rs1409706065 | 8.891E-06 | Pathogenic missense | missense_variant | 255 | R/W | 0.7596 | 0.1658 | 23.9 |
| *FFAR2* | 19:35450478:G:A | rs150050074 | 1.524E-05 | Pathogenic missense | missense_variant | 255 | R/Q | 0.6474 | 0.0396 | 23.8 |
| *FFAR2* | 19:35450507:A:G | | 1.270E-06 | Pathogenic missense | missense_variant | 265 | N/D | 0.8636 | 0.0296 | 24.3 |
| *FFAR2* | 19:35450511:C:A | | 1.270E-06 | Pathogenic missense | missense_variant | 266 | A/D | 0.9449 | 0.0946 | 23.1 |
| *FFAR2* | 19:35450514:G:A | rs775732916 | 6.351E-06 | Pathogenic missense | missense_variant | 267 | S/N | 0.7441 | 0.0243 | 18.68 |
| *FFAR2* | 19:35450520:AC:A | | 1.270E-06 | High-impact | frameshift_variant | 269 | D/X | | | |
| *FFAR2* | 19:35450522:C:A | | 1.270E-06 | Pathogenic missense | missense_variant | 270 | P/T | 0.8437 | 0.4397 | 23.5 |
| *FFAR2* | 19:35450528:C:T | | 1.270E-06 | Pathogenic missense | missense_variant | 272 | L/F | 0.582 | 0.0422 | 22.4 |
| *FFAR2* | 19:35450535:AT:A | | 1.271E-06 | High-impact | frameshift_variant | 274 | Y/X | | | |
| *FFAR2* | 19:35450540:T:C | rs1305066612 | 1.270E-06 | Pathogenic missense | missense_variant | 276 | S/P | 0.7511 | 0.025 | 24.8 |
| *FFAR2* | 19:35450653:CA:C | | 5.081E-06 | High-impact | frameshift_variant | 314 | R/X | | | |
| *FFAR2* | 19:35450692:CT:C | | 1.270E-06 | High-impact | frameshift_variant | 327 | F/X | | | |

**<u>Legend</u>:** Rare (minor allele frequency (MAF) <0.01) and pathogenic variants were identified using two classification criteria: (1) '*high-impact functional consequence'*, which includes stop gained/lost, frameshift, and splicing alterations based on Ensembl Variant Effect Predictor annotations; (2) '*pathogenic missense variants'*, which includes missense variants with pathogenic likelihood scores >0.564 from AlphaMissense, >0.025 from M-CAP (Mendelian Clinically Applicable Pathogenicity), or >20 from CADD (Combined Annotation Dependent Depletion) classifiers (highlighted in the table).

'Variant' is described as CHR:BP:OA:EA, where CHR is the chromosome position, BP is the base-pair position (GRCh38), OA is the reference allele and EA is the effect (minor) allele. MAF refers to the effect allele frequency in the UKBB cohort analysed (n=393,649), including only individuals of white-European genetic ancestry.

**GPCR:** G-protein couple receptor genes; **RPVs:** Rare pathogenic variants.

**Table S6.** SCFA-sensing GPCR rare pathogenic variants (RPVs) carriership.

| | **HTN** | | **ACS** | | **Stroke** | | **HF** | | **MACE** | |
|---|---|---|---|---|---|---|---|---|---|---|
| | controls | cases | controls | cases | controls | cases | controls | cases | controls | cases |
| **GPCR RPVs carriers** | | | | | | | | | | |
| *FFAR3* (GPR41) | 490 (0.20%) | 319 (0.21%) | 747 (0.20%) | 62 (0.22%) | 782 (0.21%) | 27 (0.22%) | 775 (0.20%) | 34 (0.24%) | 703 (0.20%) | 106 (0.23%) |
| *FFAR2* (GPR43) | 413 (0.17%) | 315 (0.21%) | 662 (0.18%) | 66 (0.23%) | 700 (0.18%) | 28 (0.23%) | 701 (0.18%) | 27 (0.19%) | 626 (0.18%) | 102 (0.22%) |
| *HCAR2* (GPR109A) | 269 (0.11%) | 204 (0.13%) | 431 (0.12%) | 42 (0.15%) | 457 (0.12%) | 16 (0.13%) | 445 (0.12%) | 28 (0.20%) | 403 (0.12%) | 70 (0.15%) |
| Any GPCR (GPR 41/43/109A) | 1167 (0.48%) | 836 (0.55%) | 1833 (0.50%) | 170 (0.59%) | 1932 (0.51%) | 71 (0.57%) | 1914 (0.50%) | 89 (0.63%) | 1725 (0.50%) | 278 (0.61%) |

**Legend**: 'GPCR RPVs carriers' is described as n (%).

**GPCR:** G-protein couple receptor genes; **HTN:** Essential hypertension; **ACS:** Acute coronary syndrome; **HF:** Heart failure; **MACE:** Major adverse cardiac events

**Table S7.** Multiple gene-based analysis: associations between RPVs in any GPCR gene (*GPR 41/43/109A*) and risk of essential hypertension and MACE.

| Phenotype | GPCR RPVs in controls (n, %) | GPCR RPVs in cases (n, %) | OR (95% CI) | P value | $P_{FDR}$ | Regression model |
|---|---|---|---|---|---|---|
| **HTN** | 1167 (0.48%) | 836 (0.55%) | **1.11 (1.01 - 1.22)** | **0.0149** | **0.0149** | model 1 |
| | | | **1.12 (1.01 - 1.23)** | **0.0142** | **0.0149** | model 2 |
| **ACS** | 1833 (0.50%) | 170 (0.59%) | 1.13 (0.96 - 1.33) | 0.0680 | 0.0828 | model 1 |
| | | | 1.13 (0.96 - 1.33) | 0.0690 | 0.0828 | model 2 |
| | | | 1.12 (0.95 - 1.33) | 0.0828 | 0.0828 | model 3 |
| **Stroke** | 1932 (0.51%) | 71 (0.57%) | 1.10 (0.86 - 1.39) | 0.2275 | 0.2277 | model 1 |
| | | | 1.11 (0.87 - 1.41) | 0.2044 | 0.2277 | model 2 |
| | | | 1.10 (0.86 - 1.39) | 0.2277 | 0.2277 | model 3 |
| **HF** | 1914 (0.50%) | 89 (0.63%) | 1.21 (0.97 - 1.50) | 0.0448 | 0.0679 | model 1 |
| | | | 1.19 (0.95 - 1.48) | 0.0641 | 0.0679 | model 2 |
| | | | 1.18 (0.95 - 1.48) | 0.0679 | 0.0679 | model 3 |
| **MACE** | 1725 (0.50%) | 278 (0.61%) | **1.19 (1.04 - 1.36)** | **0.0046** | **0.0085** | model 1 |
| | | | **1.19 (1.04 - 1.36)** | **0.0061** | **0.0085** | model 2 |
| | | | **1.18 (1.03 - 1.35)** | **0.0085** | **0.0085** | model 3 |

**Legend:** Summary statistics from multivariable logistic regression analyses testing the effect of RPV carriership status in any GPCR genes, compared between cases and controls for each phenotype of interest.

**model 1:** adjusted for age, sex, and 10 top genetic principal components.
**model 2:** adjusted for age, sex, BMI, and 10 top genetic principal components.
**model 3:** adjusted for age, sex, BMI, 10 top genetic principal components, and SMuRFs (only for MACE subtypes analyses)
**$P_{FDR}$:** FDR adjustment for models per phenotype.

**GPCR:** G-protein couple receptor genes; **RPVs:** Rare pathogenic variants; **HTN:** Essential hypertension; **ACS:** Acute coronary syndrome; **HF:** Heart failure; **MACE:** Major adverse cardiac events; **OR:** odds ratio; **CI:** confidence interval; **SMurFs:** Standard modifiable cardiovascular risk factor, including smoking, HTN, diabetes, and hypercholesterolemia.

**Table S8.** Single gene-based analysis: associations between RPVs in individual GPCR genes and risk of essential hypertension and MACE.

| Phenotype | GPCR gene | RPVs in controls (n, %) | RPVs in cases (n, %) | OR (95% CI) | P value | $P_{FDR}$ | Regression model |
|---|---|---|---|---|---|---|---|
| **HTN** | *FFAR2* | 413 (0.17%) | 315 (0.21%) | **1.20 (1.03 - 1.40)** | **0.0109** | **0.0326** | model 1 |
| | | | | **1.23 (1.05 - 1.45)** | **0.0055** | **0.0326** | model 2 |
| | *FFAR3* | 490 (0.20%) | 319 (0.21%) | 1.01 (0.87 - 1.17) | 0.4360 | 0.4990 | model 1 |
| | | | | 1.00 (0.86 - 1.17) | 0.4990 | 0.4990 | model 2 |
| | *HCAR2* | 269 (0.11%) | 204 (0.13%) | 1.14 (0.94 - 1.38) | 0.0912 | 0.1611 | model 1 |
| | | | | 1.13 (0.93 - 1.38) | 0.1074 | 0.1611 | model 2 |
| **ACS** | *FFAR2* | 662 (0.18%) | 66 (0.23%) | 1.23 (0.95 - 1.60) | 0.0577 | 0.2596 | model 1 |
| | | | | 1.25 (0.96 - 1.63) | 0.0470 | 0.2596 | model 2 |
| | | | | 1.20 (0.92 - 1.57) | 0.0877 | 0.2631 | model 3 |
| | *FFAR3* | 747 (0.20%) | 62 (0.22%) | 1.01 (0.78 - 1.32) | 0.4582 | 0.5139 | model 1 |
| | | | | 1.00 (0.76 - 1.30) | 0.5139 | 0.5139 | model 2 |
| | | | | 1.03 (0.79 - 1.36) | 0.4065 | 0.5139 | model 3 |
| | *HCAR2* | 431 (0.12%) | 42 (0.15%) | 1.16 (0.84 - 1.61) | 0.1870 | 0.3366 | model 1 |
| | | | | 1.17 (0.84 - 1.62) | 0.1783 | 0.3366 | model 2 |
| | | | | 1.13 (0.81 - 1.57) | 0.2326 | 0.3489 | model 3 |
| **Stroke** | *FFAR2* | 700 (0.18%) | 28 (0.23%) | 1.19 (0.81 - 1.74) | 0.1863 | 0.5230 | model 1 |
| | | | | 1.21 (0.82 - 1.77) | 0.1661 | 0.5230 | model 2 |
| | | | | 1.17 (0.80 - 1.72) | 0.2064 | 0.5230 | model 3 |
| | *FFAR3* | 782 (0.21%) | 27 (0.22%) | 1.05 (0.71 - 1.54) | 0.4053 | 0.5230 | model 1 |
| | | | | 1.06 (0.72 - 1.55) | 0.3930 | 0.5230 | model 2 |
| | | | | 1.08 (0.73 - 1.60) | 0.3459 | 0.5230 | model 3 |
| | *HCAR2* | 457 (0.12%) | 16 (0.13%) | 1.02 (0.62 - 1.68) | 0.4737 | 0.5230 | model 1 |
| | | | | 1.02 (0.62 - 1.69) | 0.4650 | 0.5230 | model 2 |

| | | | | | | | |
|---|---|---|---|---|---|---|---|
| | | | | 0.99 (0.60 - 1.63) | 0.5230 | 0.5230 | model 3 |
| **HF** | *FFAR2* | 701 (0.18%) | 27 (0.19%) | 0.99 (0.67 - 1.47) | 0.5102 | 0.5930 | model 1 |
| | | | | 0.98 (0.66 - 1.46) | 0.5444 | 0.5930 | model 2 |
| | | | | 0.95 (0.64 - 1.42) | 0.5930 | 0.5930 | model 3 |
| | *FFAR3* | 775 (0.20%) | 34 (0.24%) | 1.15 (0.81 - 1.62) | 0.2235 | 0.4151 | model 1 |
| | | | | 1.10 (0.77 - 1.58) | 0.2971 | 0.4456 | model 2 |
| | | | | 1.14 (0.80 - 1.64) | 0.2306 | 0.4151 | model 3 |
| | *HCAR2* | 445 (0.12%) | 28 (0.20%) | **1.62 (1.10 - 2.39)** | **0.0075** | **0.0337** | model 1 |
| | | | | **1.63 (1.10 - 2.42)** | **0.0071** | **0.0337** | model 2 |
| | | | | **1.57 (1.06 - 2.33)** | **0.0122** | **0.0365** | model 3 |
| **MACE** | *FFAR2* | 626 (0.18%) | 102 (0.22%) | 1.22 (0.98 - 1.51) | 0.0403 | 0.1059 | model 1 |
| | | | | 1.22 (0.98 - 1.52) | 0.0384 | 0.1059 | model 2 |
| | | | | 1.18 (0.94 - 1.48) | 0.0735 | 0.1186 | model 3 |
| | *FFAR3* | 703 (0.20%) | 106 (0.23%) | 1.12 (0.91 - 1.39) | 0.1418 | 0.1596 | model 1 |
| | | | | 1.10 (0.89 - 1.37) | 0.1945 | 0.1945 | model 2 |
| | | | | 1.14 (0.92 - 1.42) | 0.1192 | 0.1532 | model 3 |
| | *HCAR2* | 403 (0.12%) | 70 (0.15%) | 1.25 (0.96 - 1.63) | 0.0471 | 0.1059 | model 1 |
| | | | | 1.26 (0.96 - 1.64) | 0.0455 | 0.1059 | model 2 |
| | | | | 1.21 (0.93 - 1.59) | 0.0791 | 0.1186 | model 3 |

**Legend:** Summary statistics from multivariable logistic regression analyses testing the effect of RPV carriership status in individual GPCR genes, compared between cases and controls for each phenotype of interest. $P_{FDR:}$ FDR adjustment for multiple comparisons of four individual GPCR genes per phenotype.

**model 1:** adjusted for age, sex, and 10 top genetic principal components.
**model 2:** adjusted for age, sex, BMI, and 10 top genetic principal components.
**model 3:** adjusted for age, sex, BMI, 10 top genetic principal components, and SMuRFs (only for MACE subtypes analyses).
**$P_{FDR:}$** FDR for multiple comparisons of three individual GPCR genes and models per phenotype.

**GPCR:** G-protein couple receptor genes; **RPVs:** Rare pathogenic variants; **HTN:** Essential hypertension; **ACS:** Acute coronary syndrome; **HF:** Heart failure; **MACE:** Major adverse cardiac events; **OR:** odds ratio; **CI:** confidence interval; **SMurFs:** Standard modifiable cardiovascular risk factor, including smoking, HTN, diabetes, and hypercholesterolemia.

**Table S9.** Demographic and clinical characteristics of carriers and non-carriers of GPCR RPVs among cases (n=162,813).

| | HTN | | | ACS | | | Stroke | | | HF | | | MACE | | |
|---|---|---|---|---|---|---|---|---|---|---|---|---|---|---|---|
| | non-carriers | carriers | P value | non-carriers | carriers | P value | non-carriers | carriers | P value | non-carriers | carriers | P value | non-carriers | carriers | P value |
| N of participants | 150,929 | 836 | | 28,497 | 170 | | 12,298 | 71 | | 13,963 | 89 | | 45,233 | 278 | |
| Women (n, %) | 71,208 (47.2) | 376 (45.0) | 0.216 | 7,545 (26.5) | 43 (25.3) | 0.794 | 4,850 (39.4) | 27 (38.0) | 0.904 | 4,885 (35.0) | 30 (33.7) | 0.888 | 14,769 (32.7) | 88 (31.7) | 0.773 |
| Age, years (mean, SD) | 59.85 (6.96) | 59.89 (6.91) | 0.855 | 61.18 (6.43) | 61.22 (5.97) | 0.934 | 61.49 (6.50) | 60.52 (7.09) | 0.212 | 62.31 (6.00) | 61.87 (5.87) | 0.489 | 61.30 (6.45) | 60.98 (6.19) | 0.401 |
| BMI, kg/m$^2$ (mean, SD) | 29.05 (5.08) | 28.84 (5.06) | 0.222 | 28.90 (4.75) | 28.85 (4.56) | 0.900 | 28.61 (5.00) | 28.29 (5.25) | 0.591 | 29.81 (5.73) | 30.43 (6.00) | 0.319 | 28.92 (5.03) | 29.05 (5.15) | 0.686 |
| SBP, mmHG (mean, SD) | 149.48 (19.41) | 149.99 (20.43) | 0.461 | 144.09 (20.23) | 145.28 (21.39) | 0.457 | 145.20 (20.73) | 144.37 (20.30) | 0.748 | 145.15 (21.37) | 143.46 (21.27) | 0.462 | 144.59 (20.39) | 145.29 (21.10) | 0.577 |
| DBP, mmHg (mean, SD) | 86.08 (10.85) | 86.63 (11.27) | 0.160 | 81.80 (11.19) | 81.93 (11.02) | 0.885 | 83.18 (11.27) | 83.92 (10.15) | 0.593 | 82.06 (11.67) | 83.03 (11.23) | 0.439 | 82.40 (11.22) | 82.86 (11.04) | 0.503 |
| Smoking (current or past) (n, %) | 57,742 (38.3) | 340 (40.7) | 0.163 | 14,031 (49.2) | 94 (55.3) | 0.134 | 5,637 (45.8) | 35 (49.3) | 0.643 | 6,975 (50.0) | 42 (47.2) | 0.679 | 21,403 (47.3) | 140 (50.4) | 0.341 |

**Legend:** P value based on a t-test (for quantitative variables) or chi-square (for qualitative variables) comparing non-carriers versus carriers.

**ACS:** Acute coronary syndrome; **HTN:** Essential hypertension; **HF:** Heart failure; **MACE:** Major adverse cardiac events; **SD:** Standard deviation; **BMI:** Body mass index; **SBP:** Systolic blood pressure; **DBP:** Diastolic blood pressure.

**Table S10.** Significant comorbidity association in cases carrying GPCR RPVs.

| Comorbidity | Phenotype | Comorbidity prevalence (%) | Comorbidity in non-carriers (n, %) | Comorbidity in carriers (n, %) | OR (95% CI) * | P value | $P_{FDR}$ | Regression model |
|---|---|---|---|---|---|---|---|---|
| Cardiomyopathy (ICD-10: I42) | HTN | 1.194% | 1,789 (1.2%) | 23 (2.8%) | 2.26 (1.50 - 3.41) | $9.3 \times 10^{-5}$ | 0.0277 | model 1 |
| | | | | | 2.29 (1.52 - 3.44) | $7.8 \times 10^{-5}$ | 0.0277 | model 2 |

**Legend:** Summary statistics from multivariable logistic regression analyses testing morbidity prevalence differences between GPCR RPVs carriers and non-carriers among cases for each phenotype of interest.

**model 1:** adjusted for age and sex.
**model 2:** adjusted for age, sex, and BMI.
**$P_{FDR}$:** FDR adjustment for multiple comparisons of 297 selected ICD-10 diseases (showing a prevalence ≥1% in HTN or MACE cases) and two models (with and without BMI).

**GPCR:** G-protein couple receptor genes; **RPVs:** Rare pathogenic variants; **HTN:** Essential hypertension; **OR:** odds ratio; **CI:** confidence interval; **ICD-10:** International Classification of Disease 10th version.

**Table S11.** Demographic and clinical characteristics of carriers and non-carriers of GPCR RPVs among adequate-fiber diet consumers (n=13,858).

| | **Adequate-fiber consumers (≥25g/day for women and ≥30g/day for men)** | | |
|---|---|---|---|
| | non-carriers | carriers | P value |
| N of participants | 13,788 | 70 | |
| Women (n, %) | 9,839 (71.4) | 54 (77.1) | 0.350 |
| Age, years (mean, SD) | 57.01 (7.84) | 57.11 (7.56) | 0.911 |
| BMI, kg/m$^2$ (mean, SD) | 26.47 (4.98) | 26.96 (5.26) | 0.411 |
| SBP, mmHg (mean, SD) | 138.58 (19.77) | 140.47 (21.89) | 0.445 |
| DBP, mmHg (mean, SD) | 80.66 (10.54) | 81.08 (13.88) | 0.749 |
| Smoking (current or past) (n, %) | 3,613 (26.2) | 16 (22.9) | 0.676 |
| HTN (n, %) | **4,485 (32.5)** | **34 (48.6)** | **0.006** |
| ACS (n, %) | 689 (5.0) | 6 (8.6) | 0.275 |
| Stroke (n, %) | 316 (2.3) | 0 (0.0) | 0.379 |
| HF (n, %) | 354 (2.6) | 0 (0.0) | 0.328 |
| MACE (n, %) | 1,162 (8.4) | 6 (8.6) | 1.000 |
| Diabetes (n, %) | 989 (7.2) | 7 (10.0) | 0.496 |
| Hypercholesterolemia (n, %) | 2,070 (15.0) | 11 (15.7) | 1.000 |
| Any SMuRF (n, %) | 7,228 (52.4) | 44 (62.9) | 0.104 |

**Legend:** P value based on a t-test (for quantitative variables) or chi-square (for qualitative variables) comparing non-carriers versus carriers.

**ACS:** Acute coronary syndrome; **HTN:** Essential hypertension; **HF:** Heart failure; **MACE:** Major adverse cardiac events; **SD:** Standard deviation; **BMI:** Body mass index; **SBP:** Systolic blood pressure; **DBP:** Diastolic blood pressure; **SMurFs:** Standard modifiable cardiovascular risk factor, including smoking, HTN, diabetes, and hypercholesterolemia.

**Table S12.** Hypertension prevalence in GPCR RPV carriers versus non-carriers among adequate-fiber diet consumers (single- and multiple-gene analysis).

| **GPCR gene** | **Adequate-fiber consumers** | | **HTN prevalence in non-carriers (n, %)** | **HTN prevalence in carriers (n, %)** | **OR (95% CI)** | **P value** | **$P_{FDR}$** | **Regression model** |
|---|---|---|---|---|---|---|---|---|
| | **non-carriers** | **carriers** | | | | | | |
| **Multi-gene analysis** | | | | | | | | |
| Any GPCR (GPR 41/43/109A) | 13,788 | 70 | 4,485 (32.5%) | 34 (48.6%) | 2.14 (1.30 - 3.50) | **0.0025** | **0.0050** | model 1 |
| | | | | | 2.09 (1.25 - 3.51) | **0.0052** | **0.0052** | model 2 |
| **Single-gene analysis** | | | | | | | | |
| *FFAR2* (GPR43) | 13,832 | 26 | 4,509 (32.6%) | 10 (38.5%) | 1.39 (0.61 - 3.18) | 0.4356 | 1.0000 | model 1 |
| | | | | | 1.23 (0.52 - 2.93) | 0.6364 | 1.0000 | model 2 |
| *FFAR3* (GPR41) | 13,824 | 34 | 4,501 (32.6%) | 18 (53.9%) | 2.64 (1.30 - 5.36) | **0.0075** | **0.0229** | model 1 |
| | | | | | 2.74 (1.31 - 5.76) | **0.0076** | **0.0229** | model 2 |
| *HCAR2* (GPR109A) | 13,847 | 11 | 4,512 (32.6%) | 7 (63.6%) | 3.66 (1.03 - 12.97) | **0.0444** | 0.1333 | model 1 |
| | | | | | 3.96 (1.04 – 15.00) | **0.0430** | 0.1333 | model 2 |

**Legend:** Summary statistics from multivariable logistic regression analyses testing HTN prevalence differences between GPCR RPVs carriers and non-carriers, within a subset of adequate-fiber diet consumers (≥25g/day for women and ≥30g/day for men; n=13,858 subset from 168,142 with dietary data available).

**model 1:** adjusted for age and sex.
**model 2:** adjusted for age, sex, and BMI.
**$P_{FDR}$:** FDR adjustment for two models (with and without BMI) and three individual GPCR genes (for single-gene analysis only).

**GPCR:** G-protein couple receptor genes; **HTN:** Essential hypertension; **RPVs:** Rare pathogenic variants; **OR:** odds ratio; **CI:** confidence interval.

**Table S13.** Demographic and clinical characteristics of carriers and non-carriers of GPCR RPVs among inadequate-fiber diet consumers (n=154,284).

| | **Inadequate-fiber consumers (<25g/day for women and <30g/day for men)** | | |
|---|---|---|---|
| | non-carriers | carriers | P value |
| N of participants | 153,529 | 755 | |
| Women (n, %) | 81,324 (53.0) | 378 (50.1) | 0.119 |
| Age, years (mean, SD) | 56.36 (7.87) | 56.63 (7.89) | 0.343 |
| BMI, $kg/m^2$ (mean, SD) | 27.01 (4.57) | 27.03 (4.64) | 0.878 |
| SBP, mmHg (mean, SD) | 139.34 (19.30) | 140.58 (20.71) | 0.085 |
| DBP, mmHg (mean, SD) | 81.99 (10.55) | 82.61 (11.04) | 0.119 |
| Smoking (current or past) (n, %) | 47,308 (30.8) | 261 (34.6) | **0.029** |
| HTN (n, %) | 53,579 (34.9) | 287 (38.0) | 0.080 |
| ACS (n, %) | 9,273 (6.0) | 61 (8.1) | **0.023** |
| Stroke (n, %) | 3,627 (2.4) | 13 (1.7) | 0.300 |
| HF (n, %) | 3,920 (2.6) | 30 (4.0) | **0.019** |
| MACE (n, %) | 14,304 (9.3) | 95 (12.6) | **0.003** |
| Diabetes (n, %) | 10,858 (7.1) | 51 (6.8) | 0.789 |
| Hypercholesterolemia (n, %) | 29,394 (19.1) | 174 (23.0) | **0.008** |
| Any SMuRF (n, %) | 87,122 (56.7) | 472 (62.5) | **0.002** |

**Legend:** P value based on a t-test (for quantitative variables) or chi-square (for qualitative variables) comparing non-carriers versus carriers.

**ACS:** Acute coronary syndrome; **HTN:** Essential hypertension; **HF:** Heart failure; **MACE:** Major adverse cardiac events; **SD:** Standard deviation; **BMI:** Body mass index; **SBP:** Systolic blood pressure; **DBP:** Diastolic blood pressure; **SMurFs:** Standard modifiable cardiovascular risk factor, including smoking, HTN, diabetes, and hypercholesterolemia.

**Table S14.** Hypertension prevalence in GPCR RPV carriers versus non-carriers among inadequate-fiber diet consumers (single- and multiple-gene analysis).

| GPCR gene | Inadequate-fiber consumers | | HTN prevalence in non-carriers (n, %) | HTN prevalence in carriers (n, %) | OR (95% CI) | P value | $P_{FDR}$ | Regression model |
|---|---|---|---|---|---|---|---|---|
| | non-carriers | carriers | | | | | | |
| **Multi-gene analysis** | | | | | | | | |
| Any GPCR (GPR 41/43/109A) | 153,529 | 755 | 53,579 (34.9%) | 287 (38.0%) | 1.12 (0.96 - 1.30) | 0.1596 | 0.1595 | model 1 |
| | | | | | 1.13 (0.96 - 1.33) | 0.1393 | 0.1595 | model 2 |
| **Single-gene analysis** | | | | | | | | |
| *FFAR2* (GPR43) | 154,009 | 275 | 53,755 (34.9%) | 111 (40.4%) | 1.29 (1.00 - 1.66) | 0.0507 | 0.1522 | model 1 |
| | | | | | 1.33 (1.02 - 1.73) | **0.0342** | 0.1522 | model 2 |
| *FFAR3* (GPR41) | 153,977 | 307 | 53,755 (34.9%) | 111 (36.2%) | 1.03 (0.81 - 1.32) | 0.7961 | 1.0000 | model 1 |
| | | | | | 1.00 (0.77 - 1.30) | 0.9867 | 1.0000 | model 2 |
| *HCAR2* (GPR109A) | 154,108 | 176 | 53,801 (34.9%) | 65 (36.9%) | 0.99 (0.72 - 1.36) | 0.9403 | 1.0000 | model 1 |
| | | | | | 1.01 (0.73 - 1.41) | 0.9308 | 1.0000 | model 2 |

**Legend:** Summary statistics from multivariable logistic regression analyses testing HTN prevalence differences between GPCR RPVs carriers and non-carriers, within a subset of inadequate-fiber diet consumers (<25g/day for women and <30g/day for men; n=154,284 subset from 168,142 with dietary data available).

**model 1:** adjusted for age and sex.
**model 2:** adjusted for age, sex, and BMI.
**$P_{FDR}$:** FDR adjustment for two models (with and without BMI) and three individual GPCR genes (for single-gene analysis only).

**GPCR:** G-protein couple receptor genes; **HTN:** Essential hypertension; **RPVs:** Rare pathogenic variants; **OR:** odds ratio; **CI:** confidence interval.

**Table S15.** Demographic and clinical differences between adequate and inadequate-fiber diet consumers (n=168,142).

| | **Inadequate-fibre consumers** | **Adequate-fibre consumers** | P value |
|---|---|---|---|
| N of participants | 154,284 | 13,858 | |
| RPVs GPCR carriers (%) | 755 (0.5) | 70 (0.5) | 0.849 |
| Women (%) | 81,702 (53.0) | 9,893 (71.4) | **<0.001** |
| Age (mean, SD) | 56.36 (7.87) | 57.01 (7.84) | **<0.001** |
| BMI (mean, SD) | 27.01 (4.57) | 26.48 (4.98) | **<0.001** |
| SBP (mean, SD) | 139.34 (19.30) | 138.59 (19.78) | **<0.001** |
| DBP (mean, SD) | 82.00 (10.55) | 80.66 (10.56) | **<0.001** |
| Smoking (current or past) (n, %) | 47,569 (30.8) | 3,629 (26.2) | **<0.001** |
| HTN (n, %) | 53,866 (34.9) | 4,519 (32.6) | **<0.001** |
| ACS (n, %) | 9,334 (6.0) | 695 (5.0) | **<0.001** |
| Stroke (n, %) | 3,640 (2.4) | 316 (2.3) | 0.576 |
| HF (n, %) | 3,950 (2.6) | 354 (2.6) | 0.99 |
| MACE (n, %) | 14,399 (9.3) | 1,168 (8.4) | **<0.001** |
| Diabetes (n, %) | 10,909 (7.1) | 996 (7.2) | 0.621 |
| Hypercholesterolemia (n, %) | 29,568 (19.2) | 2,081 (15.0) | **<0.001** |
| Any SMuRF (n, %) | 87,594 (56.8) | 7,272 (52.5) | **<0.001** |

**Legend:** P value based on a t-test (for quantitative variables) or chi-square (for qualitative variables) comparing non-carriers versus carriers.

**ACS:** Acute coronary syndrome; **HTN:** Essential hypertension; **HF:** Heart failure; **MACE:** Major adverse cardiac events; **SD:** Standard deviation; **BMI:** Body mass index; **SBP:** Systolic blood pressure; **DBP:** Diastolic blood pressure; **SMurFs:** Standard modifiable cardiovascular risk factor, including smoking, HTN, diabetes, and hypercholesterolemia.
